# The DCM Project Portal: A direct-to-participant platform of The DCM Research Project

**DOI:** 10.1101/2023.06.22.23291764

**Authors:** Elizabeth S. Jordan, Phoenix L. Grover, Jay Lin, Carl A. Starkey, Elizabeth A. Finley, Hanyu Ni, Ray E. Hershberger

## Abstract

**Study Objective:** To develop a digital platform to conduct family-based, dilated cardiomyopathy (DCM) genetic research.

**Design:** Innovative approaches are needed to achieve large family enrollment targets. The DCM Project Portal, a direct-to-participant electronic recruitment, consent, and communication tool, was designed using prior experience with traditional enrollment methods, characteristics and feedback of current participants, and internet access of the US population.

**Participants:** DCM patients (probands) and their family members.

**Results:** The portal was designed as a self-guided, three module (registration, eligibility, and consent) process with internally created supporting informational and messaging resources integrated throughout. The experience can be tailored to user type and the format adapted with programmatic growth. Characteristics of participants of the recently completed DCM Precision Medicine Study were assessed as an exemplary user population. A majority of the diverse (34% non-Hispanic Black (NHE-B), 9.1% Hispanic; 53.6% female) proband (n=1223) and family members (n=1781) participants aged ≥18 years reported *not at all* or *rarely* having problems learning about their health from written information (81%) and a high confidence in completing medical forms (77.2% *very much* or *often* confident). A majority of participants across age and race-ethnicity groups reported internet access, with highest rates of no reported access in those ≥77 years, NHE-B, and Hispanic, which reflects patterns similar to rates reported by the US Census Bureau as of 2021.

**Conclusions:** Digital enrollment tools offer opportunity to improve access and efficiency. The portal is an example of a digital approach to family-based genetic research.

## 1. Introduction

Clinical genetics research can be conducted in case-based designs, where the unit of investigation is a patient with a phenotype of interest. This model has been fruitful for gene discovery and genotype-phenotype correlations, and particularly for high penetrance genes and variants. However, in complex cardiovascular genetic disease, case-only approaches may be insufficient. Shifting the paradigm to viewing a family as the unit of investigation, i.e., the patient with the phenotype of interest *and* their relatives, has promise to provide a more comprehensive view of shared rare and common variant, epigenic, environmental, and other factors within a family that contribute to the marked clinical and genetic complexity in heritable cardiovascular disease.^1^ Although a family-based approach is recommended in the clinical genetic evaluation of cardiomyopathy,^2^ recruiting a family unit is labor-intensive and challenging, and persists as a major barrier to implementation of a family-based research design.

The Dilated Cardiomyopathy (DCM) Research Project is a family-based research program established in the 1990s that aims to investigate the genetics of DCM, a condition characterized by left ventricular enlargement and systolic dysfunction and a leading cause of heart failure.^1, 3^ When the cause of DCM remains unknown after a thorough clinical evaluation, an underlying genetic cause is suspected, but clinical genetic testing identifies a cause in only about one-third of cases.^4, 5^ The unsolved clinical and genetic complexity of DCM in patients and their family members drives DCM Research Project investigations.

The DCM Precision Medicine study, a recently completed multi-site, National Institutes of Health-funded study conducted by the DCM Research Project, enrolled >1200 DCM patients (probands) and more than 2000 of their family members from diverse racial and ethnic backgrounds to perform clinical DCM screening and genetic testing.^6–8^ The aims of the DCM Precision Medicine study were to test the hypothesis that DCM has substantial genetic basis and to evaluate the effectiveness of a family communication intervention in improving the uptake of family member clinical screening.^7, 8^ Data from family units has been critical to the emerging findings from the Precision Medicine study,^6, 9,10^ but despite these initial informative reports and others from this study,^11, 12^ many questions remain. To continue to decipher the complexity of DCM genetics, an essential step toward advancing preventive and therapeutic approaches to genetic DCM, participation of a much larger number of families from diverse backgrounds is needed.^13–15^ The DCM Research Project aims to achieve this through the aggregation of a much larger cohort of DCM families, nominally 10,000, for the DCM Discovery Study.

The DCM Precision Medicine study used a traditional, in-person enrollment model to conduct a family-based study design,^8^ and while successful, was labor intensive. To achieve the Discovery Study’s ambitious enrollment target, novel approaches are required. With the emergence of electronic health portals and applications, successful implementation of digital recruitment and electronic data collection in numerous cardiovascular clinical trials,^16, 17^ and nearly 93% of American adults reporting access to internet connected devices as of 2021^18^ (Figure 1; Supplementary Table S1), there is substantial opportunity to leverage web-based methods. A move to web-based approaches was rated favorably by a majority of sampled DCM Research Project participants and researchers.^19^

**Figure 1.**
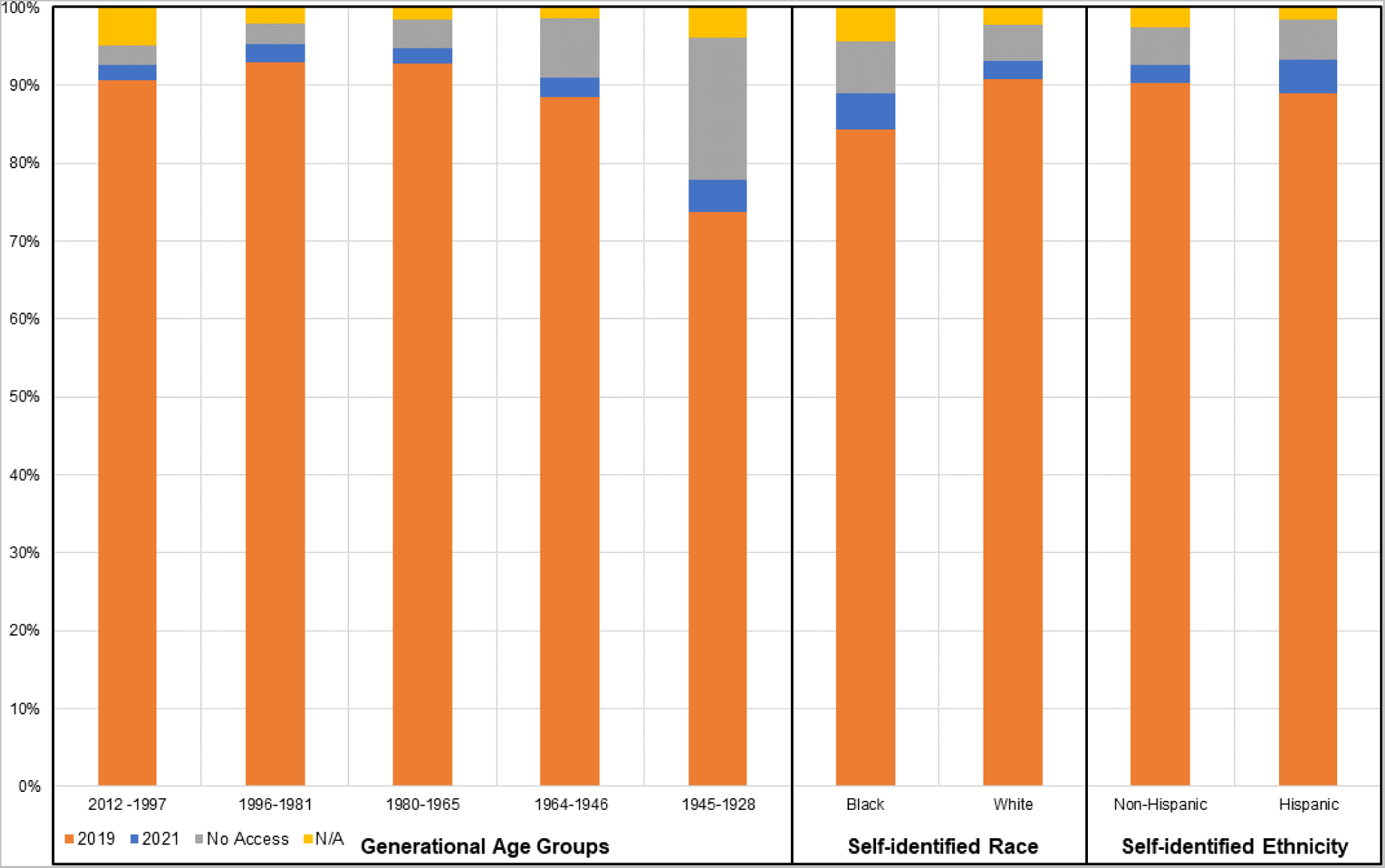
Reported internet access in the US population in 2019 and 2021 according to US Census Data The percentage of individuals in the population within generational age groups by year of birth, self-identified race, and self-identified ethnicity that reported having access to internet at home according to The United States Census Bureau. Individuals who reported having access to the internet, with or without paying a cell phone company or internet service provider, in 2019 (orange) are shown with the increase for each subgroup from 2019 to 2021 shown in blue. The years 2019 and 2021 were selected to demonstrate any increase in access before and after the COVID-19 pandemic, which increased across all groups. The percentage reporting to have no access at home as of 2021 is shown in gray and those living in group quarters or housing units vacant at the time of data collection in 2021 are shown in yellow. Generational age groups including generations with individuals ≥18 years of age as of 2023 are presented, including Generation Z (born 2012-1997), Millennials (1996-81), Generation X (1980-65), Baby Boomer (1964-46), and Silent Generation (1945-28). The data included in this figure are also provided in a table format in Supplementary Table S2.

The DCM Project Portal is a direct-to-participant electronic recruitment, consent, and communication tool designed for family recruitment to the DCM Research Project. The objectives of the portal are to maximize staff efficiency and improve access to participation in The DCM Discovery study, while also facilitating long-term participant engagement. Here, we describe the design and rationale of the DCM Project Portal to conduct family-based genetic research.

## 2. Materials and Methods

### 2.1 Design of The DCM Project Portal

The key needs for the design of an electronic approach to recruitment and enrollment were derived from the study’s extensive experience with family-based study designs using a traditional, in-person process in prior DCM Research Project investigations (section 2.3). Most important has been the relationships built with participating families thus far, which have been central to study success over time. Thus, the central goal for the portal was to serve as a communication interface and to continue to support these important relationships with participating families by providing an open, accessible, and easy communication platform. Other data also contributed to the key needs of the Portal design, including characteristics of the DCM Precision Medicine study participants, US Census-derived estimates of internet access of the US population, and current participant feedback regarding important aspects of web-based models. The following features were prioritized.

The first key feature was a “direct-to-participant” design, meaning that the web-based process can be entirely participant guided. This was prioritized to reduce staff time needed to recruit and enroll each individual in a family unit as well as to increase the accessibility of participation beyond only those that receive care at DCM Consortium sites (Supplementary Figure S1). A direct-to-participant approach is also an important strategy to distribute self-administered methods of data collection.

A second feature important for the portal was to be adaptable to continued growth of the research program and to be able to be tailored to individual user’s needs. We anticipate new ancillary studies, greater numbers of requests for families to invite additional relatives to join the study, and greatly expanded data collection needs. To accommodate continued programmatic growth, an adaptable structure allowing for modules or features to be easily added will yield the most benefit and allow the portal to be a long-term resource. For families, similar adaptability is needed at the user level, so that the user experience can be tailored to the needs of participants, such as activation of proband- or family member-participant specific items or allowing users to skip items that do not pertain for a more efficient experience.

A third key design feature was to foster engagement of participants with the research program, congruent with the central goal of maintaining long-term relationships with participating families. By engagement we primarily mean maintaining participant-study staff contact to so that participants are able to keep study staff informed of relevant changes to their history. Engagement also includes remaining available, for example, to receive communications regarding new studies they may be eligible for or new data that may need to be collected.

Other important features were focused on operational aspects and prioritized feedback from current study participants, including an advised time to complete the process of 30 minutes or less, having technical support available, and the ability to accommodate varying skillsets.^19^ The characteristics of current DCM Precision Medicine study participants were considered as a model participant demographic for user populations for future studies to anticipate potential support and infrastructure needs. Informational text and videos were internally developed by DCM Research Project investigators and personnel.

A three-phase beta testing process was completed (Supplementary Figure S2). Technical development of The DCM Project Portal was conducted by an external developer under the direction of study personnel. The portal is securely housed at The Ohio Supercomputer Center (OSC) (www.osc.edu) and requires two-factor authentication for login.

### 2.2 Demographic, Personal Health Literacy, and Internet Usage Information

Demographic, personal health literacy, and internet usage information was collected from The DCM Precision Medicine study participants at the time of enrollment. These data were used to consider the design and potential efficacy for a web-based model. Structured interviews collected social and demographic information. Personal health literacy and internet usage characteristics were assessed by six questions in a self-administered survey, inquiring difficulty, confidence, and assistance needs with understanding and completing medical information about themselves in addition to internet accessibility and use for medical purposes. Ages groups were selected based on year of birth by generational classifications. Race-ethnicity was based on self-identification and grouped as non-Hispanic Black, non-Hispanic White, and Hispanic.

To evaluate population internet access, responses to the “Access to Internet” variable, inquiring if individuals have access to internet connected devices at home by paid or unpaid services, was exported from the Public Use Microdata Sample (PUMS) using the US Census Bureau’s Microdata Access Tool (MDAT) based on self-identified race, ethnicity, and year of birth by generational classifications. ACS 1-Year Estimates PUMS was selected to generate the data.

### 2.3 Design considerations from The DCM Precision Medicine Study experience

The DCM Precision Medicine study was a multi-site, cross-sectional family-based study executed through the multi-site DCM Consortium led by The Ohio State University (OSU) (Supplementary Figure S1).^8^ Institutional Review Boards at OSU and all clinical sites approved the study in the initial period. During the study, a single IRB at the University of Pennsylvania was created with reliance agreements from all clinical sites. Written informed consent was obtained from all participants. The single IRB at the University of Pennsylvania has approved the DCM Project Portal.

Using an in-person model, the study accrued a diverse (nearly half non-Hispanic Black and nearly half female) including over 1200 probands with idiopathic DCM (clinical criteria provided in Supplemental Material) between June 2016-March 2020 and over 2000 of their family members through March 2021. Family member recruitment was a multi-step process and required several individuals, staff and participants, as well as substantial oversight to achieve. In short, enrolled probands were asked to refer first degree relatives (parents, siblings, children) and other family member(s) affected with heart disease to the study. With written permission, study staff worked with probands to follow up on referrals and enroll relatives. Data was collected for all participants by medical record review and questionnaires collected by structured interview regarding cardiovascular history, quality of life, health behaviors, family dynamics and family medical history. Of note, enrollment activity was truncated in part by the COVID-19 pandemic due to the limitations on in-person research activities.

## 3. Results

### 3.1 The Participant Experience

The portal is available to previously consented and newly interested individuals. Newly interested individuals may self-discover the study or be referred by a provider or other individual familiar with the research program. A workflow outlining the three-step process tailored to the user type is shown in Figure 2, including (1) Registration, (2) Eligibility, and (3) Consent Modules. Supporting text (“*Read More*”) and video (“*Watch More*”) resources are available for each item throughout. By current participant advisement,^23^ users are able to message study staff (“*Ask More*”) at any point. The process is customizable by entering unique, staff-generated codes to allow skipping of sections based on user type, such as indicating that the user has already had eligibility screening, provided consent, or is a family member of a current participant. Upon completion of the Consent Module, the user is shown a full copy of the consent form for final review in REDCap,^26^ and an electronic signature is collected. A PDF file of the signed form is immediately available for download upon submitting. Electronically formatted surveys and medical record release forms are available once consented, reducing staff time that would typically be required to collect this information.

**Figure 2.**
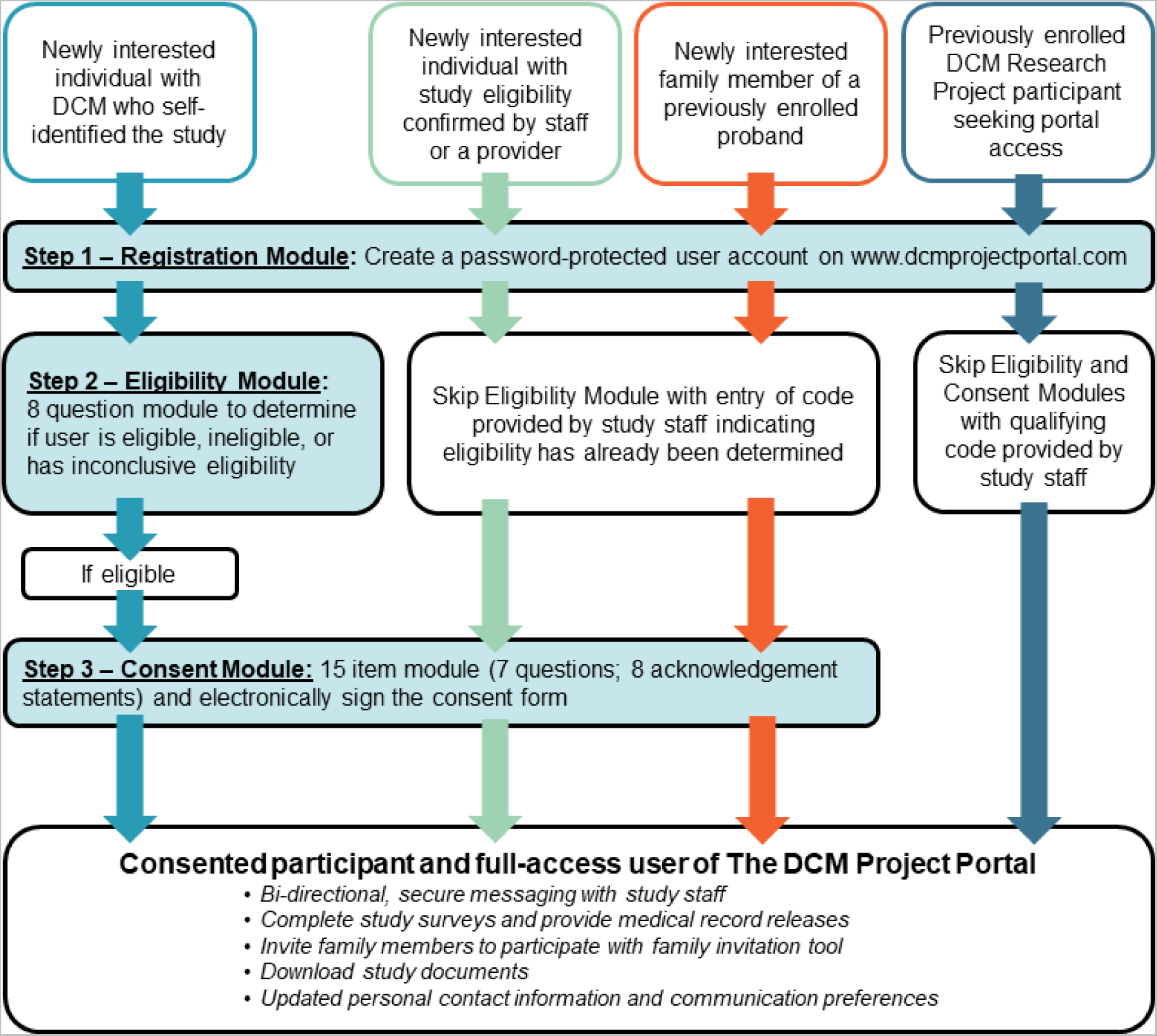
The Three Step DCM Project Portal Process by User Type The three-step process tailored to the user type is shown, including (1) Registration, (2) Eligibility, and (3) Consent Modules. The Registration step establishes an account by collecting basic demographic and contact information, permission to communicate through text messaging, and preferred language. To complete Registration, users are required to create a password so that they may exit and return to their account. If the registered user does not provide skip code indicating that they are eligible, they are directed to the Eligibility Module. This section consists of a series of eight self-guided “yes,” “no,” or “not sure” questions adapted from the study’s eligibility criteria (Supplementary Material). Upon completion, logic is built to determine in real time if a user is eligible, ineligible, or has inconclusive eligibility, which requires additional staff review to determine the status. The Consent Module for eligible individuals is similarly built, consisting of a 15-item design with seven questions and eight acknowledgment statements. Upon completion of the Consent Module an electronic signature is collected. Once a user has completed the process specific to their user type, as indicated by the four colored pathways shown, they have full access to the user features of the portal.

The Consent Module was designed to prioritize key components of the informed consent process, including accommodation of the dynamic nature of this process and maximizing the autonomy of the research volunteer.^20–22^ The simple question-based design, provision of *Read, Watch, and Ask More* resources, and the option to save, exit, and return to modules were put in place to ensure users are adequately informed, not coerced, and understand their choice to participate.

### 3.2 Participant engagement tools

The *Family Invitation* tool is a participant-directed family recruitment tool, where an individual can send a personally signed, pre-written invitation email explaining the study and how to get involved. This digital family recruitment process can occur without staff involvement unless assistance is requested, in contrast to the described previous time intensive process (section 2.3). In addition, secure messaging is available for users once registered. When a new message is sent, participants receive a text (or email as preferred) notifying them that a message is available, resembling an electronic health record communication model.

### 3.3 The Staff Interface

The landing page of the staff interface shows a live log of user activity. Staff can access records, correspond, review and update eligibility and consent statuses, download documents, and view if family members have been invited at an individual user level. As noted, staff can produce unique codes to validate a user’s consent or eligibility status, as applicable, to tailor the user experience. Study surveys and newsletters can be distributed at individual or group-level communications. Communication fields are also built in for staff-to-staff information exchange.

### 3.4 Participant Characteristics Portal Development

Characteristics of participants aged 18 years and older at the time of enrollment to the DCM Precision Medicine Study was evaluated as an exemplary population of anticipated portal users (Table 1). This included 1223 probands and 1781 first-degree relatives (FDRs) 43.7% and 60.4% were female, respectively; 42.2% probands and 28.3% of family members identified as non-Hispanic Black, 49.3% and 62.3% non-Hispanic white, and 8.5% and 9.4% Hispanic. Additional characteristics are summarized in Table 1.

**Table 1.** Characteristics of participants age eighteen and older in the DCM Precision Medicine Study by Participant Type

| Characteristic | Participant Type - No. (%) |  |  |
| --- | --- | --- | --- |
|  | Total | Proband | Family Member |
| **Total** | 3004 (100%) | 1223 (100%) | 1781 (100%) |
| **Sex** |  |  |  |
| Female | 1610 (53.6%) | 535 (43.7%) | 1075 (60.4%) |
| Male | 1394 (46.4%) | 688 (56.3%) | 706 (46.4%) |
| **Self-reported Race-Ethnicity Group** |  |  |  |
| Non-Hispanic Black | 1014 (34.0%) | 516 (42.2%) | 498 (28.3%) |
| Non-Hispanic White | 1697 (56.9 %) | 602 (49.3%) | 1095 (62.3%) |
| Hispanic | 270 (9.1%) | 104 (8.5%) | 166 (9.4%) |
| **Age at enrollment, y** |  |  |  |
| 18-26 | 347 (11.6%) | 55 (4.5%) | 292 (16.6%) |
| 27-42 | 794 (26.6%) | 262 (21.4%) | 532 (30.2%) |
| 43-58 | 988 (33.1%) | 516 (42.2%) | 472 (26.8%) |
| 59-76 | 777 (26.1%) | 368 (30.1%) | 409 (23.3%) |
| ≥77 | 75 (2.5%) | 21 (1.7%) | 54 (3.1%) |
| **Age at DCM diagnosis, y<sup>a</sup>** | N/A<sup>a</sup> | 44.2±13.5y (4.7 - 82.7) | N/A<sup>a</sup> |
| **Geographic Location** |  |  |  |
| Northeast | 348 (11.6%) | 153 (12.5%) | 195 (11.0%) |
| Midwest | 1040 (34.7%) | 390 (31.9%) | 650 (36.7%) |
| South | 1101 (36.8%) | 493 (40.3%) | 608 (34.3%) |
| West | 505 (16.9%) | 186 (15.2%) | 319 (18.0%) |
| **Education, y** |  |  |  |
| ≤12 | 1076 (37.8%) | 451 (39.3%) | 625 (36.8%) |
| >12 | 1773 (62.2%) | 698 (60.7%) | 1075 (63.2%) |
| **Work Status** |  |  |  |
| Employed for wages | 1380 (47.1%) | 396 (33.3%) | 984 (56.4%) |
| Self-employed | 278 (9.5%) | 115 (9.7%) | 163 (9.3%) |
| Out of work | 271 (9.2%) | 187 (15.7%) | 84 (4.8%) |
| A home-maker | 133 (4.5%) | 49 (4.1%) | 84 (4.8%) |
| A student | 108 (3.7%) | 23 (1.9%) | 85 (4.9%) |
| Retired | 664 (22.6%) | 353 (29.7%) | 311 (17.8%) |
| **Marital Status** |  |  |  |
| Married | 1510 (51.5%) | 650 (54.7%) | 860 (49.3%) |
| Divorced | 347 (11.8%) | 164 (13.8%) | 183 (10.5%) |
| Widowed | 114 (3.9%) | 48 (4.0%) | 66 (3.8%) |
| Separated | 65 (2.2%) | 34 (2.9%) | 31 (1.8%) |
| Never Married | 713 (24.3%) | 224 (18.8%) | 489 (28.0%) |
| Member of an unmarried couple | 130 (4.4%) | 46 (3.9%) | 84 (4.8%) |

<sup>a</sup>age at diagnosis only calculated for probands as many family members were not affected with dilated cardiomyopathy at the time of enrollment

A majority of participants across all age and race-ethnicity groups reported minimal difficulty (*not at all* or *rarely* having problems) learning about their medical condition due to understanding written information and high confidence in filling out medical forms independently (*very much* or *often* feeling confident) (Table 2). While a majority of participants reported having internet access at home, 31.9% of participants ≥77 years old reported no access (Table 2), though this age group only represented 2.5% of the DCM Precision Medicine study (Table 1). Other groups with highest reports of no home internet access in included participants who self-identified as non-Hispanic Black or Hispanic ethnicity (Table 2), reflecting similar access patterns to that of the general population (Figure 1). Additional personal health literacy and internet usage characteristics of participants by age group and self-identified race-ethnicity are shown in Table 2.

**Table 2.** Personal health literacy and internet access and usage characteristics of DCM Precision Medicine Study Participants by age and race-ethnicity groups

| Questions | Responses | <u>Generational Age Groups - No. (%)</u> |  |  |  |  |  | <u>Self-reported Race-Ethnicity Groups<sup>a</sup> - No. (%)</u> |  |  |  |
| --- | --- | --- | --- | --- | --- | --- | --- | --- | --- | --- | --- |
|  |  | Total<sup>b</sup> (N) | 18-26 | 27-42 | 43-58 | 59-76 | ≥77y | Total<sup>b</sup> | NHE-B<sup>a</sup> | NHE-W<sup>a</sup> | Hispanic |
| **Personal Health Literacy** |  | N=2933 | N=342 | N=787 | N=974 | N=758 | N=72 | N=2910 | N=973 | N=1671 | N=266 |
| *How often do you have problems learning about your medical condition because of difficulty understanding written information?* | Not at all | 1562 | 158 (46.2%) | 444 (56.4%) | 542 (55.6%) | 383 (50.5%) | 35 (48.6%) | 1551 | 539 (55.4%) | 879 (52.6%) | 133 (50.0%) |
|  | Rarely | 814 | 104 (30.4%) | 203 (25.8%) | 259 (26.6%) | 227 (29.9%) | 21 (29.2%) | 804 | 257 (26.4%) | 483 (28.9%) | 64 (24.1%) |
|  | Some | 376 | 54 (15.8%) | 100 (12.7%) | 112 (11.5%) | 102 (13.5%) | 8 (11.1%) | 374 | 102 (10.5%) | 224 (13.4%) | 48 (18.0%) |
|  | Often | 67 | 9 (2.6%) | 16 (2.0%) | 24 (2.5%) | 13 (1.7%) | 5 (6.9%) | 67 | 25 (2.6%) | 35 (2.1%) | 7 (2.6%) |
|  | Very Much | 43 | 7 (2.0%) | 12 (1.5%) | 13 (1.3%) | 9 (1.2%) | 2 (2.8%) | 43 | 20 (2.1%) | 16 (1.0%) | 7 (2.6%) |
| *How confident are you filling out medical forms by yourself?* | Not at all | 179 | 10 (2.9%) | 35 (4.4%) | 59 (6.1%) | 60 (7.9%) | 15 (20.8%) | 179 | 69 (7.1%) | 81 (4.8%) | 29 (10.9%) |
|  | Rarely | 109 | 19 (5.6%) | 26 (3.3%) | 41 (4.2%) | 19 (2.5%) | 4 (5.6%) | 108 | 44 (4.5%) | 52 (3.1%) | 12 (4.5%) |
|  | Some | 315 | 64 (18.7%) | 62 (7.9%) | 87 (8.9%) | 88 (11.6%) | 14 (19.4%) | 314 | 87 (8.9%) | 197 (11.8%) | 30 (11.3%) |
|  | Often | 466 | 107 (31.3%) | 112 (14.2%) | 131 (13.4%) | 103 (13.6%) | 13 (18.1%) | 463 | 131 (13.5%) | 279 (16.7%) | 53 (19.9%) |
|  | Very Much | 1798 | 131 (38.3%) | 541 (68.7%) | 635 (65.2%) | 465 (61.3%) | 26 (36.1%) | 1780 | 617 (63.4%) | 1028 (61.5%) | 135 (50.8%) |
| *How often do you have someone help you read hospital materials?* | Not at all | 1607 | 133 (38.9) | 483 (61.4%) | 566 (58.1%) | 399 (52.6%) | 26 (36.1%) | 1589 | 550 (56.5%) | 923 (55.2%) | 116 (43.6%) |
|  | Rarely | 660 | 107 (31.3%) | 180 (22.9%) | 190 (19.5%) | 163 (21.5%) | 20 (27.8%) | 658 | 203 (20.9%) | 394 (23.6%) | 61 (22.9%) |
|  | Some | 337 | 52 (15.2%) | 74 (9.4%) | 109 (11.2%) | 92 (12.1%) | 10 (13.9%) | 336 | 95 (9.8%) | 199 (11.9%) | 42 (15.8%) |
|  | Often | 147 | 28 (8.2%) | 23 (2.9%) | 45 (4.6%) | 45 (5.9%) | 6 (8.3%) | 145 | 50 (5.1%) | 70 (4.2%) | 25 (9.4%) |
|  | Very Much | 104 | 9 (2.6%) | 16 (2.0%) | 37 (3.8%) | 34 (4.5%) | 8 (11.1%) | 104 | 40 (4.1%) | 50 (3.0%) | 14 (5.3%) |
| **Internet Access and Usage** |  | N=2933 | N=342 | N=787 | N=974 | N=758 | N=72 | N=2910 | N=973 | N=1671 | N=266 |
| *Do you have internet access at home?* | Yes<sup>c</sup> | 2422 | 279 (81.6%) | 661 (84.0%) | 810 (83.2%) | 625 (82.5%) | 47 (65.3%) | 2400 | 705 (72.5%) | 1499 (89.7%) | 196 (73.7%) |
|  | No | 445 | 55 (16.1%) | 115 (14.6%) | 143 (14.7%) | 109 (14.4%) | 23 (31.9%) | 444 | 245 (25.2%) | 138 (8.3%) | 61 (22.9%) |
| *In the past 12 months, did you use the internet, whether from home or somewhere else to* | Yes | 2213 | 239 (69.9%) | 630 (80.1%) | 744 (76.4%) | 561 (74.0%) | 39 (54.2%) | 2196 | 695 (71.4%) | 1326 (79.4%) | 175 (65.8%) |
|  | No | 632 | 94 (27.5%) | 142 (18.0%) | 199 (20.4%) | 167 (22.0%) | 30 (41.7%) | 626 | 241 (24.8%) | 307 (18.4%) | 78 (29.3%) |
| *look for health or medical information for yourself?* |  |  |  |  |  |  |  |  |  |  |  |
| *In the past 12 months, how often did you use the internet to look for health or medical information for yourself or someone else?* | Not Applicable | 13 | 3 (0.9%) | 0 | 3 (0.3%) | 5 (0.7%) | 2 (2.8%) | 13 | 4 (0.4%) | 9 (0.5%) | 0 (0.0%) |
|  | Once a week | 635 | 58 (17.0%) | 196 (24.9%) | 226 (23.2%) | 146 (19.3%) | 9 (12.5%) | 630 | 236 (24.3%) | 321 (19.2%) | 73 (27.4%) |
|  | Once a month | 633 | 69 (20.2%) | 180 (22.9%) | 228 (23.4%) | 151 (19.9%) | 5 (6.9%) | 629 | 198 (20.3%) | 385 (23.0%) | 46 (17.3%) |
|  | Every few months | 693 | 77 (22.5%) | 200 (25.4%) | 222 (22.8%) | 179 (23.6%) | 15 (20.8%) | 685 | 194 (19.9%) | 439 (26.3%) | 52 (19.5%) |
|  | Less often than every few months | 850 | 123 (36.0%) | 194 (24.7%) | 267 (27.4%) | 233 (30.7%) | 33 (45.8%) | 844 | 300 (30.8%) | 466 (27.9%) | 78 (29.3%) |

<sup>a</sup>NHE-B=non-Hispanic Black; NHE-W=non-Hispanic White
<sup>b</sup>The number of total responses differs from the total counts of individuals in each demographic group shown in table 1 due to missing responses.
<sup>c</sup>Individuals who marked “yes” to having internet access at home include those who had access through their phone, dialup, and/or broadband or highspeed internet.

## 4. Discussion

The DCM Project Portal to our knowledge is the first multi-faceted, web-based tool developed to support family-based genetic investigations of DCM. As described, the Portal is designed to facilitate the enrollment of large numbers of patients with DCM and their family members and has been approved by the DCM Research Project’s single IRB. Beta testing has been accomplished and the Portal has now entered service.

Family units are a critical scientific need for investigations of The DCM Research Project. However, working with families adds to recruitment complexities. It has been observed across families with shared genetic disease risk that information sharing can be selective^23–25^ and several barriers to communicating about genetic risk information have been described in families, including emotional, geographic, health literacy, self-efficacy, guilt, and others, all of which can make recruitment of a family unit, rather than an individual patient, more challenging.^23, 24^ Incomplete participation of family members at risk for genetic cardiovascular disease has been observed by The DCM Research Project^10^ and others.^26, 27^ Offering multiple modalities and simple processes to engage one’s family has the potential to help bypass communication barriers and improve participation of family units.

Direct-to-participant digital recruitment and enrollment modalities are becoming increasingly used to conduct clinical research. As the population internet usage continues to increase (Figure 1; Supplementary Tables S1-S5), the integration of electronic strategies will be needed to achieve large enrollment goals, with potential to capture more individuals and maximize efficiency of study staff time. However, the development of digital platforms requires extensive effort and must be tailored to unique needs of a specific study or research program.^16^

Diversity in genetic and genomic research is essential, and the use of multiple enrollment modalities may also be critical to support diversity of participants.^28^ Although US Census data suggests internet access has increased since 2019 across generational age and race-ethnicity groups (Figure 1), possibly in part due to the shift toward digital environments as a result of the COVID-19 pandemic, individuals in the population who are self-identified black, Hispanic, and were born between the years of 1964-1946 (Baby boomer generation) or 1928-1945 (Silent Generation), had the highest reported rates of not having access to the internet at their home in 2021 (6.6%, 5.1%, and 7.6%, 18.2% respectively; Figure 1; Supplementary Tables S1-S5). A similar pattern as was reported by current participants identifying as non-Hispanic Black, Hispanic, and over the age of 77 years (Table 2). The rates of no access were higher in participants in these groups compared to the proportions reported in the population, however data were collected from study participants at the time of enrollment which could be as early as 2016 when connectivity rates would have likely been lower in general. To remedy this issue and to maximize access and support participants of diverse backgrounds web-and non-web options need to be made available, as has also been shown by others^29^ and emphasized by study participants.^19^

While initial beta testing has been completed (Supplementary Figure S2), opportunities for improvement are anticipated. Specifically, we will monitor the self-guided, question based Eligibility Module performance, as when previously surveyed current participants and researchers were asked, “*How confident are you in [your/your patient’s] ability to accurately answer questions about your eligibility for The DCM Research Project by answering a series of questions about your cardiovascular health history*,” only 56% of DCM Project researchers were confident or somewhat confident participants could do so, contrasting with a high level of confidence (87% confident or somewhat confident) reported by the sampled participants.^19^ A similar high confidence level and lack of difficulty in understanding and reporting one’s medical information was also reported by the larger set of DCM Precision Medicine Study participants (Table 2). The supportive *Read More, Watch More,* and *Ask More* features were developed to help mitigate this potential challenge that will be closely evaluated with increasing participant accrual.

An additional area for monitoring is potential risk for loss to follow up with a web-initiated approach, which could limit the development of a personal relationship with a participant. As an initial preventive measure, automated message reminders connected with text message alerts are issued if modules are incomplete or if they have a new message. The text-based communication eliminates the participant need to independently remember to return to the portal by actively engaging with them. Text messaging approaches have also been successful in other trials in improving medication adherence.^30^ Other strategies may be required if loss to follow up is observed at an undesirable rate.

Beyond anticipated challenges, we look forward to areas for future innovation, such as a method for administration of randomized interventions. Having participants consented for recontact and electronically accessible also enables easy-access and low-pressure recruitment pathways to new study and trial opportunities. Although optimization of digital approaches can be tedious to develop and maintain, the potential positive impact on participant access and efficiency of research operations presents substantial opportunity for studies seeking large sample sizes and long-term engagement.

## Conclusions

The DCM Project Portal is a web-based, direct-to-participant platform tailored to the DCM Research Project. A similar web-based design may be useful for other clinical genetics and family-based studies. Customization of the framework reported herein to other programs with similar needs has the potential to augment traditional processes.

## Data Availability

All data produced in the present study are available upon reasonable request to the authors.

## Acknowledgements

The investigators thank the participating families who have participated in The DCM Research Project program, without whom this effort would not be possible. We also thank the Ohio Supercomputer Center and Switchbox Inc. for their contributions to and support of this project.

## Supplemental Material

### Inclusion/Exclusion Criteria for probands in the DCM Precision Medicine Study

All probands have met diagnostic criteria for idiopathic DCM: left ventricular ejection fraction <50%; left ventricular enlargement (echo-derived left ventricular end-diastolic dimension ≥95th percentile for gender/height). In addition, they were in one of the target ethnicity-ancestry groups, were able to communicate in English (or in Spanish at sites approved to recruit individuals of Hispanic ethnicity), were able to give informed consent (or assent and parental consent for children), and were willing and able to participate in a family-based study. Relatives also were expected to satisfy these same requirements except for meeting diagnostic criteria for idiopathic DCM. Risk factors considered conventional for DCM, such as obesity, routinely treated hypertension, alcohol use/abuse, peripartum cardiomyopathy, or the presence of left-ventricular non-compaction, were not used as exclusion criteria.

Exclusion criteria included: Coronary artery disease (CAD) causing ischemic cardiomyopathy (>50% narrowing, any major epicardial coronary artery; clinical testing is indicated to exclude CAD routinely in (1) males >40 years or females >45 years without risk factors and (2) males or females >30 years with risk factors); Primary valvular disease; Cardiotoxic drug exposure, including Adriamycin and other cancer chemotherapeutics; Other forms of cardiomyopathy (e.g., hypertrophic cardiomyopathy, arrhythmogenic right ventricular cardiomyopathy, restrictive cardiomyopathy, Chagas cardiomyopathy); Congenital/structural heart disease; Sarcoid; Amyloid; Iron overload; Other active multisystem disease that may plausibly cause DCM (e.g., hypereosinophilic syndrome, cardiac involvement with connective tissue disease, Loeffler’s endocarditis, endomyocardial fibrosis); Severe and untreated or untreatable hypertension (defined as systolic blood pressures routinely >180 mm Hg and/or diastolic blood pressures >120 mm Hg. Untreated hypertension is an individual receiving no medications, and untreatable hypertension is an individual with severe hypertension persisting despite multidrug regimens and/or associated with other multisystem disease (e.g., scleroderma, other vasculitides, etc)); Exclusion beyond a reasonable doubt of all other detectable causes of cardiomyopathy (other than genetic) at the time of primary DCM diagnosis using a rigorous clinical standard congruent with an expert approach rendered by a board-certified heart failure/transplant cardiologist.

### Tables S1-S5. Reported access to the internet in the United States population by generational age group and self-identified race and Hispanic ethnicity

To estimate internet connectivity in the population by self-identified race, ethnicity, generational age groups, data was exported from the Public Use Microdata Sample (PUMS) using the US Census Bureau’s Microdata Access Tool (MDAT). ACS 1-Year Estimates PUMS was selected with the corresponding year to generate the dataset. Tables were generated by selecting the “Access to Internet” variable and the aforementioned demographic variables. “Age” was customized to captured to encompass age groups defined by birth years within generations (Generation Z: 1997-2012, Millennials: 1981-86, Generation X: 1965-80, Baby Boomers: 1946-64, Silent Generation: 1928-45, WWII generation: 1901-1927). Data were exported for each variable combination for both 2019 and 2021 to determine if there was a reported increase between the two years as a possible impact of the COVID-19 pandemic on internet connectivity.

**Table S1.** Total ACS 1-Year Estimates Access to the Internet in 2019 and 2021 According to the Public Use Microdata Sample.

| **Access to the Internet in the United States (2019)** |  |  |
| --- | --- | --- |
|  | 2019 | 2021 |
| *N/A<sup>a</sup>* | 8084362 (2.5%) | 7760859 (2.3%) |
| *Yes, Total (with or without paying a service<sup>b</sup>)* | 295581562 (90.0%) | 308066247 (92.8%) |
| *Yes, by paying a service* | 288656692 (87.9%) | 301737502 (90.9%) |
| *Yes, without paying a service* | 6924870 (2.1%) | 6328745 (1.9%) |
| *No access<sup>c</sup>* | 24573599 (7.5%) | 16066639 (4.8%) |
| **Total** | **328239523 (100%)** | **331893745 (100%)** |

<sup>a</sup>N/A refers to individuals who lived in group quarters or housing units that were vacant at the time of data collection
<sup>b</sup>service refers to paying a cell phone company or internet service provider
<sup>c</sup>No responses are regarding to access at home; may not be representative of access to public internet sources

**Table S2.** Reported Internet Access in the US Population in 2019 and 2021 According to Census Data.

|  | 2019 (%) |  |  |  |  | 2021 (%) |  |  |  |  | Change in Reported Internet Access between 2019 and 2021 |
| --- | --- | --- | --- | --- | --- | --- | --- | --- | --- | --- | --- |
|  | Yes, Total (with or without paying a service) | Yes, by paying a service<sup>b</sup> | Yes, without paying a service | No Access<sup>c</sup> | N/A<sup>d</sup> | Yes, Total (with or without paying a service) | Yes, by paying a service<sup>b</sup> | Yes, without paying a service | No Access<sup>c</sup> | N/A<sup>d</sup> |  |
| **Generational Age Groups<sup>a</sup>**<br>(generation name, corresponding years of birth listed) |  |  |  |  |  |  |  |  |  |  |  |
| Generation Z (2012-1997) | 90.69% | 88.53% | 2.16% | 4.54% | 4.77% | 92.68% | 90.74% | 1.94% | 2.41% | 4.91% | +1.99% |
| Millennials (1996-1981) | 93.03% | 91.02% | 2.01% | 4.67% | 2.30% | 95.28% | 93.62% | 1.65% | 2.68% | 2.04% | +2.24% |
| Generation X (1980-1965) | 92.88% | 91.03% | 1.85% | 5.49% | 1.63% | 94.88% | 93.16% | 1.72% | 3.60% | 1.52% | +2.00% |
| Baby Boomer (1964-1946) | 88.58% | 86.46% | 2.12% | 9.96% | 1.46% | 91.06% | 88.98% | 2.08% | 7.56% | 1.38% | +2.48% |
| Silent Generation (1945-1928) | 73.72% | 70.72% | 2.99% | 22.75% | 3.53% | 77.91% | 74.79% | 3.12% | 18.16% | 3.92% | +4.20% |
| **Self-identified Race** |  |  |  |  |  |  |  |  |  |  |  |
| Black | 84.35% | 81.44% | 2.91% | 11.36% | 4.29% | 89.03% | 86.54% | 2.50% | 6.64% | 4.32% | +4.68% |
| White | 90.80% | 88.77% | 2.03% | 6.99% | 2.21% | 93.09% | 91.30% | 1.78% | 4.66% | 2.25% | +2.29% |
| **Self-identified Ethnicity** |  |  |  |  |  |  |  |  |  |  |  |
| Non-Hispanic | 90.28% | 88.22% | 2.06% | 7.12% | 2.60% | 92.71% | 90.86% | 1.86% | 4.78% | 2.51% | +2.43% |
| Hispanic | 89.02% | 86.69% | 2.33% | 9.12% | 1.87% | 93.28% | 91.16% | 2.12% | 5.11% | 1.61% | +4.26% |

<sup>a</sup>only showing generations including those aged ≥18 years as of 2023 sorted by generational name and years of birth.
<sup>b</sup>service refers to paying a cell phone company of internet service provider
<sup>c</sup>No responses are regarding to access at home; may not be representative of access to public internet sources
<sup>d</sup>N/A refers to individuals who lived in group quarters or housing units that were vacant at the time of data collection

**Table S3a.** ACS 1-Year Estimates Access to the Internet in 2019 According to the Public Use Microdata Sample by Generational Age Group.

| 2019 Internet Usage by Generational Cohorts |  |  |  |  |  |
| --- | --- | --- | --- | --- | --- |
| Source: ACS 1-Year Estimates Public Use Microdata Sample 2019 |  |  |  |  |  |
| Weight used: PWGTP |  |  |  |  |  |
| Access to the Internet (ACCESS) |  |  |  |  |  |
| Generational Age Groups<sup>a</sup> (generation name, corresponding years of birth listed) | Total | N/A<sup>b</sup> | Yes, by paying a service<sup>c</sup> | Yes, without paying a service | No access<sup>d</sup> |
| **Total<sup>a</sup>** | 328239523 | 8084362 (2.46%) | 288656692 (87.94%) | 6924870 (2.11%) | 24573599 (7.49%) |
| **Generation Z (2012-1997)** | 67805245 | 3231114 (4.77%) | 60026497 (88.53%) | 1465901 (2.16%) | 3081733 (4.54%) |
| **Millennials (1996-1981)** | 71525288 | 1641951 (2.30%) | 65104337 (91.02%) | 1438565 (2.01%) | 3340435 (4.67%) |
| **Generation X (1980-1965)** | 65308606 | 1063134 (1.63%) | 59447159 (91.03%) | 1210772 (1.85%) | 3587541 (5.49%) |
| **Baby Boomer (1964-1946)** | 71803346 | 1047398 (1.46%) | 62078234 (86.46%) | 1523892 (2.12%) | 7153822 (9.96%) |
| **Silent Generation (1945-1928)** | 22850856 | 806891 (3.53%) | 16160835 (70.72%) | 683925 (2.99%) | 5199205 (22.75%) |
| **WWII (1901-1924)<sup>e</sup>** | 1880765 | 278439 (14.8%) | 862494 (45.86%) | 85710 (4.56%) | 654122 (34.78%) |

<sup>a</sup>only showing generations including those aged $\geq 18$ years as of 2023 sorted by generational name and years of birth. Total also includes the younger Generation Alpha which does not include individuals aged $\geq 18$ years as of 2023.
<sup>b</sup>N/A refers to individuals who lived in group quarters or housing units that were vacant at the time of data collection
<sup>c</sup>service refers to paying a cell phone company of internet service provider
<sup>d</sup>No responses are regarding to access at home; may not be representative of access to public internet sources
<sup>e</sup>99 is the upper age limit of the Census data and not reflective of the actual age cutoff of this generation

**Table S3b.** ACS 1-Year Estimates Access to the Internet in 2021 According to the Public Use Microdata Sample by Generational Age Group.

| 2021 Internet Usage by Generational Cohorts |  |  |  |  |  |
| --- | --- | --- | --- | --- | --- |
| Source: ACS 1-Year Estimates Public Use Microdata Sample 2021 |  |  |  |  |  |
| Weight used: PWGTP |  |  |  |  |  |
| Access to the Internet (ACCESSINET) |  |  |  |  |  |
| Generational Age Groups<sup>a</sup><br>(generation name, corresponding<br>years of birth listed) | Total | N/A<sup>b</sup> | Yes, by paying a service<sup>c</sup> | Yes, without paying a<br>service | No access<sup>d</sup> |
| **Total<sup>a</sup>** | 331893745 | 7760859 (2.34%) | 301737502 (90.91%) | 6328745 (1.91%) | 16066639 (4.84%) |
| **Generation Z (2012-1997)** | 69075253 | 3394180 (4.91%) | 62680388 (90.74%) | 1339025 (1.94%) | 1661660 (2.41%) |
| **Millennials (1996-1981)** | 72109485 | 1471669 (2.04%) | 67512342 (93.62%) | 1191720 (1.65%) | 1933754 (2.68%) |
| **Generation X (1980-1965)** | 65748113 | 1000636 (1.52%) | 61252802 (93.16%) | 1127977 (1.72%) | 2366698 (3.60%) |
| **Baby Boomer (1964-1946)** | 70571030 | 975402 (1.38%) | 62796417 (88.98%) | 1466078 (2.08%) | 5333133 (7.56%) |
| **Silent Generation (1945-1928)** | 18692329 | 733138 (3.92%) | 13980278 (74.79%) | 583471 (3.12%) | 3395442 (18.16%) |
| **WWII (1901-1924)<sup>e</sup>** | 1168943 | 164797 (14.1%) | 611421 (52.31%) | 57470 (4.92%) | 335255 (28.68%) |

<sup>a</sup>only showing generations including those aged $\geq 18$ years as of 2023 sorted by generational name and years of birth. Total also includes the younger Generation Alpha which does not include individuals aged $\geq 18$ years as of 2023.
<sup>b</sup>N/A refers to individuals who lived in group quarters or housing units that were vacant at the time of data collection
<sup>c</sup>service refers to paying a cell phone company or internet service provider
<sup>d</sup>No responses are regarding to access at home; may not be representative of access to public internet sources
<sup>e</sup>99 is the upper age limit of the Census data and not reflective of the actual age cutoff of this generation

**Table S4a.** ACS 1-Year Estimates Access to the Internet in 2019 According to the Public Use Microdata Sample by Self-identified Race.

| 2019 Internet Usage by Race |  |  |  |  |  |
| --- | --- | --- | --- | --- | --- |
| Source: ACS 1-Year Estimates Public Use Microdata Sample 2019 |  |  |  |  |  |
| Weight used: PWGTP |  |  |  |  |  |
| Access to the Internet (ACCESS) |  |  |  |  |  |
| Recoded detailed race code | Total | N/A<sup>a</sup> | Yes, by paying a service<sup>b</sup> | Yes, without paying a service | No access<sup>c</sup> |
| Total | 328239523 | 8084362 (2.46%) | 288656692 (87.94%) | 6924870 (2.11%) | 24573599 (7.49%) |
| White | 236532194 | 5233723 (2.21%) | 209980189 (88.77%) | 4794213 (2.03%) | 16524069 (6.99%) |
| Black or African American | 42043031 | 1802226 (4.29%) | 34241467 (81.44%) | 1223914 (2.91%) | 4775424 (11.36%) |
| American Indian | 2248236 | 66416 (2.95%) | 1718308 (76.43%) | 64888 (2.89%) | 398624 (17.73%) |
| Alaska Native | 112956 | 3611 (3.20%) | 90278 (79.92%) | 1199 (1.06%) | 17868 (15.82%) |
| American Indian and Alaska Native tribes specified; or American Indian or Alaska Native, not specified and no other races | 480428 | 29074 (6.05%) | 363711 (75.71%) | 11995 (2.50%) | 75648 (15.75%) |
| Asian | 18608918 | 369549 (1.99%) | 17384042 (93.42%) | 243401 (1.31%) | 611926 (3.29%) |
| Native Hawaiian and Other Pacific Islander | 610822 | 21488 (3.52%) | 520773 (85.26%) | 12008 (1.97%) | 56553 (9.26%) |
| Some other race | 16266146 | 284217 (1.75%) | 14005776 (86.1%) | 367539 (2.26%) | 1608614 (9.89%) |
| Two or More Races | 11336792 | 274058 (2.42%) | 10352148 (91.31%) | 205713 (1.81%) | 504873 (4.45%) |

<sup>a</sup>N/A refers to individuals who lived in group quarters or housing units that were vacant at the time of data collection
<sup>b</sup>service refers to paying a cell phone company of internet service provider
<sup>c</sup>No responses are regarding to access at home; may not be representative of access to public internet sources

**Table S4b.** ACS 1-Year Estimates Access to the Internet in 2021 According to the Public Use Microdata Sample by Self-identified Race.

| 2021 Internet Usage by Race |  |  |  |  |  |
| --- | --- | --- | --- | --- | --- |
| Source: ACS 1-Year Estimates Public Use Microdata Sample 2021 |  |  |  |  |  |
| Weight used: PWGTP |  |  |  |  |  |
| Access to the Internet (ACCESSINET) |  |  |  |  |  |
| Recoded detailed race code | Total | N/A<sup>a</sup> | Yes, by paying a service<sup>b</sup> | Yes, without paying a service | No access<sup>c</sup> |
| Total | 331893745 | 7760859 (2.34%) | 301737502 (90.91%) | 6328745 (1.91%) | 16066639 (4.84%) |
| White | 202954389 | 4570818 (2.25%) | 185306842 (91.30%) | 3620575 (1.78%) | 9456154 (4.66%) |
| Black or African American | 40217512 | 1739001 (4.32%) | 34802991 (86.54%) | 1003596 (3.50%) | 2671924 (6.64%) |
| American Indian | 2562058 | 44465 (1.74%) | 2193725 (85.62%) | 81593 (3.18%) | 242275 (9.46%) |
| Alaska Native | 124111 | 5162 (4.16%) | 102668 (82.72%) | 3110 (2.51%) | 13171 (10.61%) |
| American Indian and Alaska Native tribes specified; or American Indian or Alaska Native, not specified and no other races | 475644 | 37044 (7.79%) | 384921 (80.93%) | 13027 (2.74%) | 40652 (8.55%) |
| Asian alone | 19174507 | 330048 (1.72%) | 18196497 (94.90%) | 236779 (1.23%) | 411183 (2.14%) |
| Native Hawaiian and Other Pacific Islander | 623315 | 17319 (2.78%) | 556979 (89.36%) | 20497 (3.29%) | 28520 (4.58%) |
| Some other race | 23899490 | 330223 (1.38%) | 21666481 (90.66%) | 548625 (2.30%) | 1354161 (5.67%) |
| Two or More Races | 41862719 | 686779 (1.64%) | 38526398 (92.03%) | 800943 (1.91%) | 1848599 (4.42%) |

<sup>a</sup>N/A refers to individuals who lived in group quarters or housing units that were vacant at the time of data collection
<sup>b</sup>service refers to paying a cell phone company of internet service provider
<sup>c</sup>No responses are regarding to access at home; may not be representative of access to public internet sources

**Table S5a.**
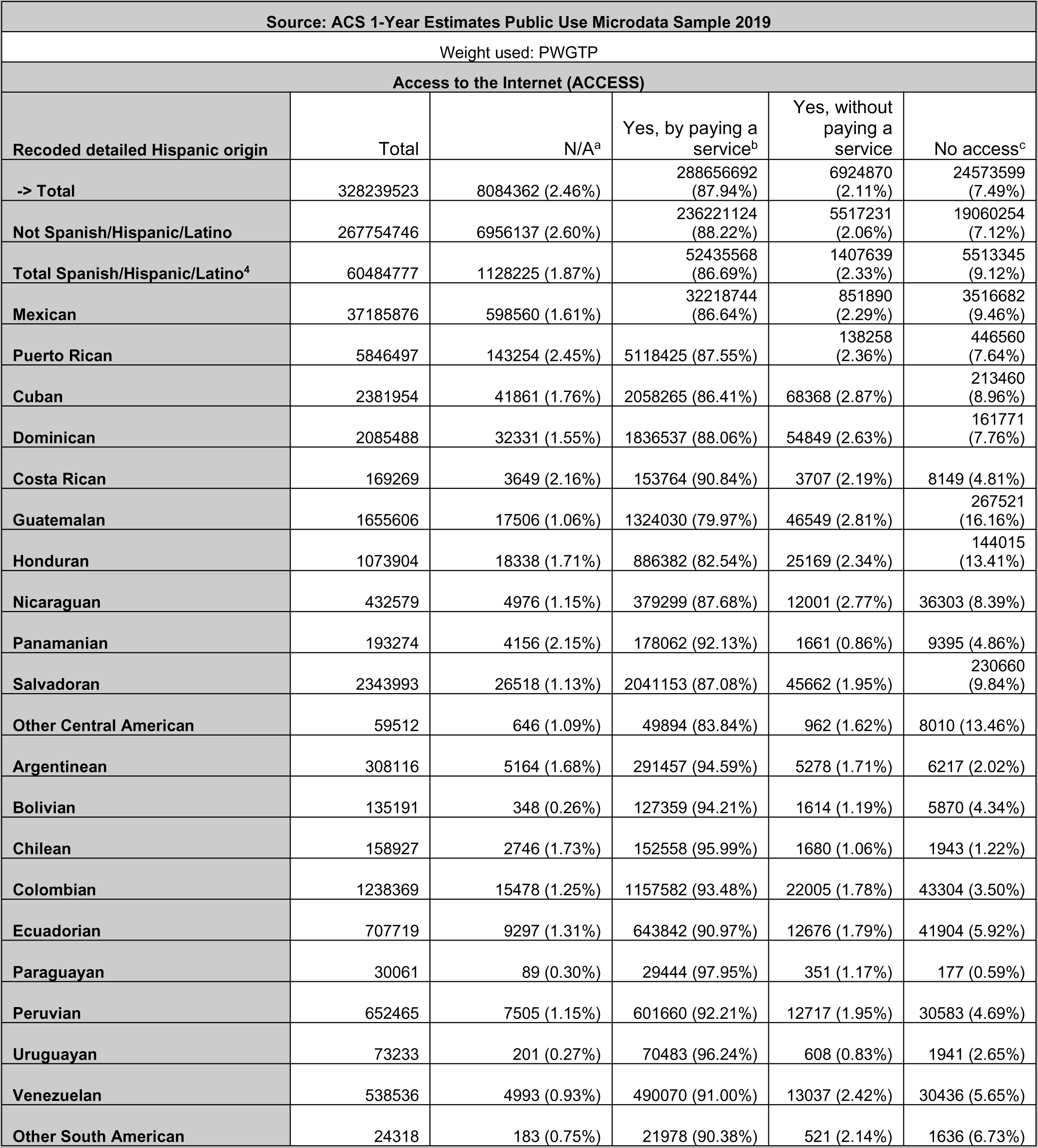
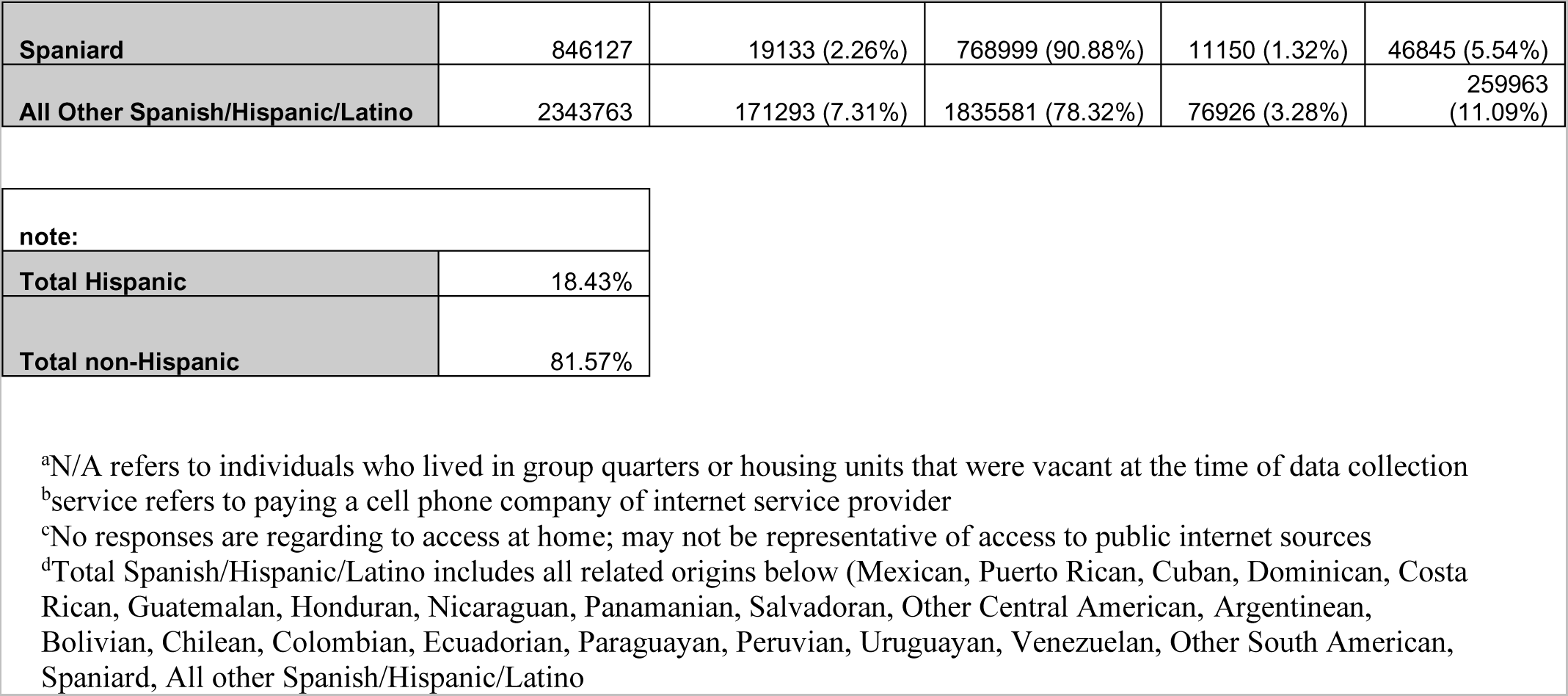
ACS 1-Year Estimates Access to the Internet in 2019 According to the Public Use Microdata Sample by Hispanic Ethnicity.

| Source: ACS 1-Year Estimates Public Use Microdata Sample 2019 |  |  |  |  |  |
| --- | --- | --- | --- | --- | --- |
| Weight used: PWGTP |  |  |  |  |  |
| Access to the Internet (ACCESS) |  |  |  |  |  |
| Recoded detailed Hispanic origin | Total | N/A<sup>a</sup> | Yes, by paying a service<sup>b</sup> | Yes, without paying a service | No access<sup>c</sup> |
| -> Total | 328239523 | 8084362 (2.46%) | 288656692 (87.94%) | 6924870 (2.11%) | 24573599 (7.49%) |
| Not Spanish/Hispanic/Latino | 267754746 | 6956137 (2.60%) | 236221124 (88.22%) | 5517231 (2.06%) | 19060254 (7.12%) |
| Total Spanish/Hispanic/Latino<sup>d</sup> | 60484777 | 1128225 (1.87%) | 52435568 (86.69%) | 1407639 (2.33%) | 5513345 (9.12%) |
| Mexican | 37185876 | 598560 (1.61%) | 32218744 (86.64%) | 851890 (2.29%) | 3516682 (9.46%) |
| Puerto Rican | 5846497 | 143254 (2.45%) | 5118425 (87.55%) | 138258 (2.36%) | 446560 (7.64%) |
| Cuban | 2381954 | 41861 (1.76%) | 2058265 (86.41%) | 68368 (2.87%) | 213460 (8.96%) |
| Dominican | 2085488 | 32331 (1.55%) | 1836537 (88.06%) | 54849 (2.63%) | 161771 (7.76%) |
| Costa Rican | 169269 | 3649 (2.16%) | 153764 (90.84%) | 3707 (2.19%) | 8149 (4.81%) |
| Guatemalan | 1655606 | 17506 (1.06%) | 1324030 (79.97%) | 46549 (2.81%) | 267521 (16.16%) |
| Honduran | 1073904 | 18338 (1.71%) | 886382 (82.54%) | 25169 (2.34%) | 144015 (13.41%) |
| Nicaraguan | 432579 | 4976 (1.15%) | 379299 (87.68%) | 12001 (2.77%) | 36303 (8.39%) |
| Panamanian | 193274 | 4156 (2.15%) | 178062 (92.13%) | 1661 (0.86%) | 9395 (4.86%) |
| Salvadoran | 2343993 | 26518 (1.13%) | 2041153 (87.08%) | 45662 (1.95%) | 230660 (9.84%) |
| Other Central American | 59512 | 646 (1.09%) | 49894 (83.84%) | 962 (1.62%) | 8010 (13.46%) |
| Argentinean | 308116 | 5164 (1.68%) | 291457 (94.59%) | 5278 (1.71%) | 6217 (2.02%) |
| Bolivian | 135191 | 348 (0.26%) | 127359 (94.21%) | 1614 (1.19%) | 5870 (4.34%) |
| Chilean | 158927 | 2746 (1.73%) | 152558 (95.99%) | 1680 (1.06%) | 1943 (1.22%) |
| Colombian | 1238369 | 15478 (1.25%) | 1157582 (93.48%) | 22005 (1.78%) | 43304 (3.50%) |
| Ecuadorian | 707719 | 9297 (1.31%) | 643842 (90.97%) | 12676 (1.79%) | 41904 (5.92%) |
| Paraguayan | 30061 | 89 (0.30%) | 29444 (97.95%) | 351 (1.17%) | 177 (0.59%) |
| Peruvian | 652465 | 7505 (1.15%) | 601660 (92.21%) | 12717 (1.95%) | 30583 (4.69%) |
| Uruguayan | 73233 | 201 (0.27%) | 70483 (96.24%) | 608 (0.83%) | 1941 (2.65%) |
| Venezuelan | 538536 | 4993 (0.93%) | 490070 (91.00%) | 13037 (2.42%) | 30436 (5.65%) |
| Other South American | 24318 | 183 (0.75%) | 21978 (90.38%) | 521 (2.14%) | 1636 (6.73%) |
| **Spaniard** | 846127 | 19133 (2.26%) | 768999 (90.88%) | 11150 (1.32%) | 46845 (5.54%) |
| **All Other Spanish/Hispanic/Latino** | 2343763 | 171293 (7.31%) | 1835581 (78.32%) | 76926 (3.28%) | 259963 (11.09%) |

|  |  |
| --- | --- |
| **note:** |  |
| **Total Hispanic** | 18.43% |
| **Total non-Hispanic** | 81.57% |

<sup>a</sup>N/A refers to individuals who lived in group quarters or housing units that were vacant at the time of data collection
<sup>b</sup>service refers to paying a cell phone company or internet service provider
<sup>c</sup>No responses are regarding to access at home; may not be representative of access to public internet sources
<sup>d</sup>Total Spanish/Hispanic/Latino includes all related origins below (Mexican, Puerto Rican, Cuban, Dominican, Costa Rican, Guatemalan, Honduran, Nicaraguan, Panamanian, Salvadoran, Other Central American, Argentinean, Bolivian, Chilean, Colombian, Ecuadorian, Paraguayan, Peruvian, Uruguayan, Venezuelan, Other South American, Spaniard, All other Spanish/Hispanic/Latino)

**Table S5b.**
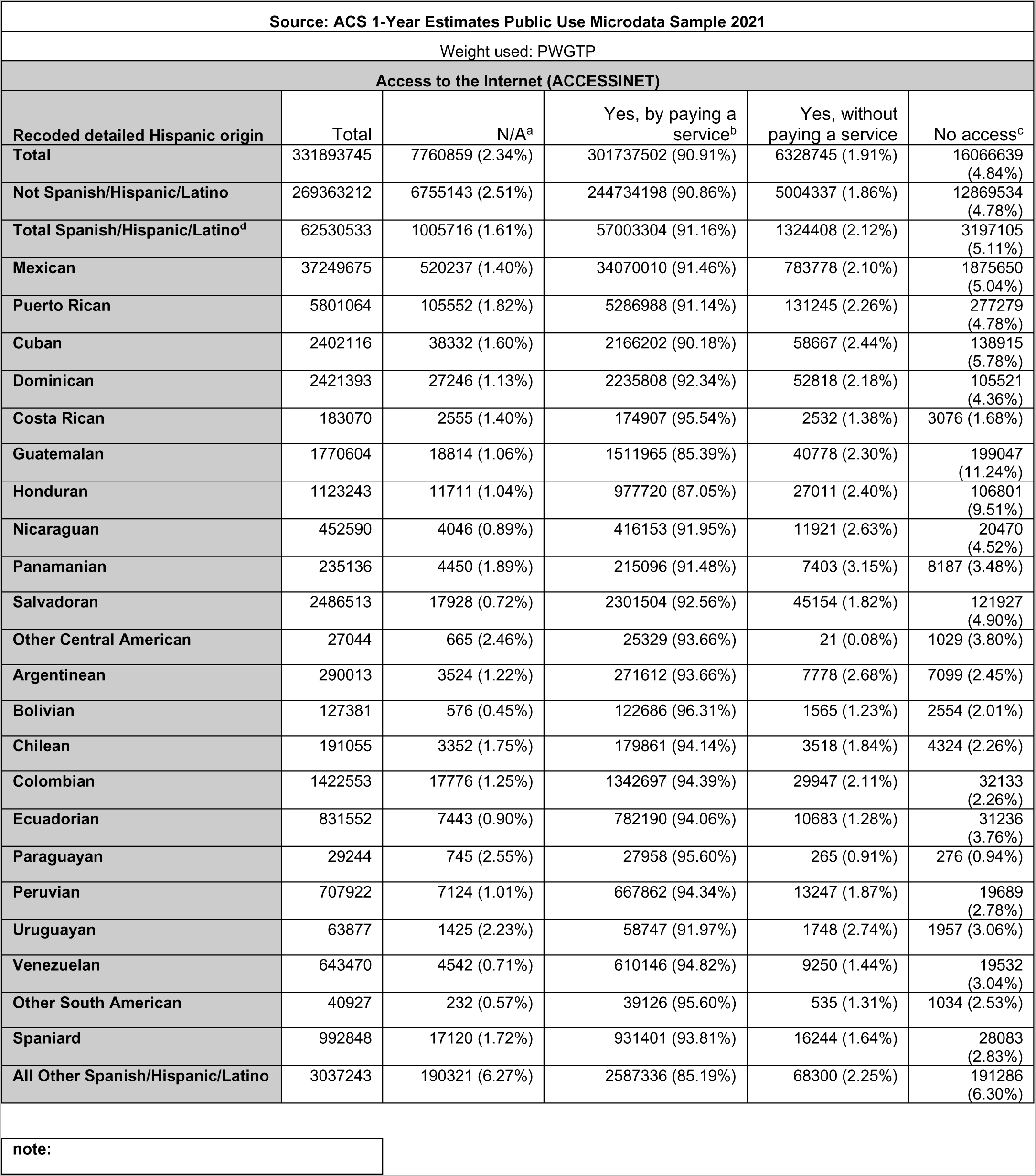
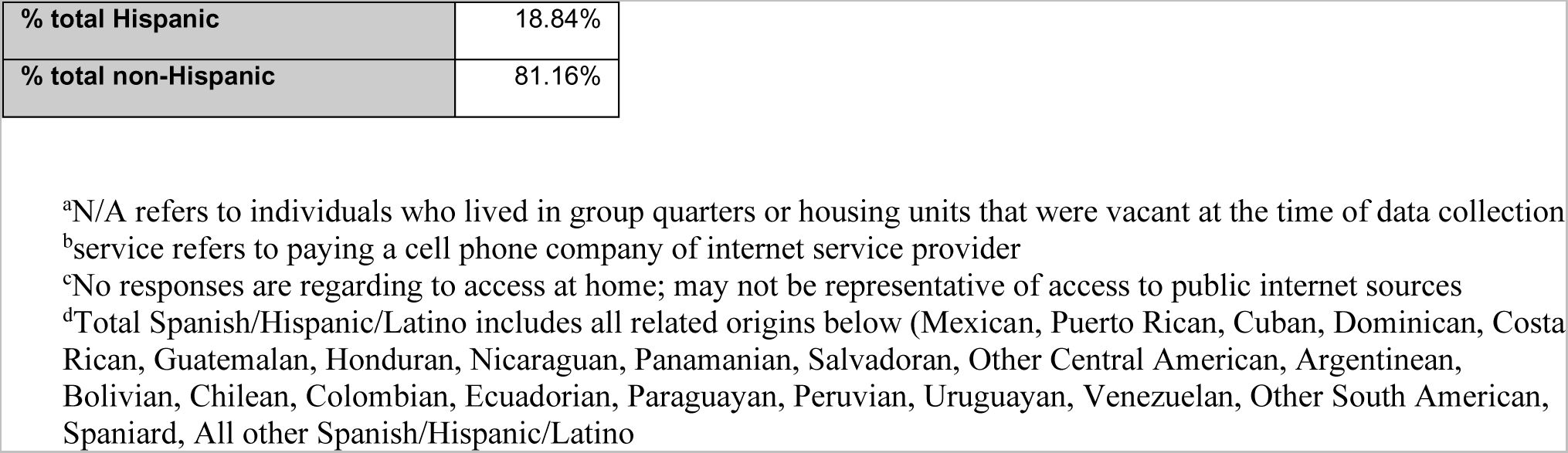
ACS 1-Year Estimates Access to the Internet in 2021 According to the Public Use Microdata Sample by Hispanic Ethnicity.

| Source: ACS 1-Year Estimates Public Use Microdata Sample 2021 |  |  |  |  |  |
| --- | --- | --- | --- | --- | --- |
| Weight used: PWGTP |  |  |  |  |  |
| Access to the Internet (ACCESSINET) |  |  |  |  |  |
| Recoded detailed Hispanic origin | Total | N/A<sup>a</sup> | Yes, by paying a service<sup>b</sup> | Yes, without paying a service | No access<sup>c</sup> |
| **Total** | 331893745 | 7760859 (2.34%) | 301737502 (90.91%) | 6328745 (1.91%) | 16066639 (4.84%) |
| **Not Spanish/Hispanic/Latino** | 269363212 | 6755143 (2.51%) | 244734198 (90.86%) | 5004337 (1.86%) | 12869534 (4.78%) |
| **Total Spanish/Hispanic/Latino<sup>d</sup>** | 62530533 | 1005716 (1.61%) | 57003304 (91.16%) | 1324408 (2.12%) | 3197105 (5.11%) |
| **Mexican** | 37249675 | 520237 (1.40%) | 34070010 (91.46%) | 783778 (2.10%) | 1875650 (5.04%) |
| **Puerto Rican** | 5801064 | 105552 (1.82%) | 5286988 (91.14%) | 131245 (2.26%) | 277279 (4.78%) |
| **Cuban** | 2402116 | 38332 (1.60%) | 2166202 (90.18%) | 58667 (2.44%) | 138915 (5.78%) |
| **Dominican** | 2421393 | 27246 (1.13%) | 2235808 (92.34%) | 52818 (2.18%) | 105521 (4.36%) |
| **Costa Rican** | 183070 | 2555 (1.40%) | 174907 (95.54%) | 2532 (1.38%) | 3076 (1.68%) |
| **Guatemalan** | 1770604 | 18814 (1.06%) | 1511965 (85.39%) | 40778 (2.30%) | 199047 (11.24%) |
| **Honduran** | 1123243 | 11711 (1.04%) | 977720 (87.05%) | 27011 (2.40%) | 106801 (9.51%) |
| **Nicaraguan** | 452590 | 4046 (0.89%) | 416153 (91.95%) | 11921 (2.63%) | 20470 (4.52%) |
| **Panamanian** | 235136 | 4450 (1.89%) | 215096 (91.48%) | 7403 (3.15%) | 8187 (3.48%) |
| **Salvadoran** | 2486513 | 17928 (0.72%) | 2301504 (92.56%) | 45154 (1.82%) | 121927 (4.90%) |
| **Other Central American** | 27044 | 665 (2.46%) | 25329 (93.66%) | 21 (0.08%) | 1029 (3.80%) |
| **Argentinean** | 290013 | 3524 (1.22%) | 271612 (93.66%) | 7778 (2.68%) | 7099 (2.45%) |
| **Bolivian** | 127381 | 576 (0.45%) | 122686 (96.31%) | 1565 (1.23%) | 2554 (2.01%) |
| **Chilean** | 191055 | 3352 (1.75%) | 179861 (94.14%) | 3518 (1.84%) | 4324 (2.26%) |
| **Colombian** | 1422553 | 17776 (1.25%) | 1342697 (94.39%) | 29947 (2.11%) | 32133 (2.26%) |
| **Ecuadorian** | 831552 | 7443 (0.90%) | 782190 (94.06%) | 10683 (1.28%) | 31236 (3.76%) |
| **Paraguayan** | 29244 | 745 (2.55%) | 27958 (95.60%) | 265 (0.91%) | 276 (0.94%) |
| **Peruvian** | 707922 | 7124 (1.01%) | 667862 (94.34%) | 13247 (1.87%) | 19689 (2.78%) |
| **Uruguayan** | 63877 | 1425 (2.23%) | 58747 (91.97%) | 1748 (2.74%) | 1957 (3.06%) |
| **Venezuelan** | 643470 | 4542 (0.71%) | 610146 (94.82%) | 9250 (1.44%) | 19532 (3.04%) |
| **Other South American** | 40927 | 232 (0.57%) | 39126 (95.60%) | 535 (1.31%) | 1034 (2.53%) |
| **Spaniard** | 992848 | 17120 (1.72%) | 931401 (93.81%) | 16244 (1.64%) | 28083 (2.83%) |
| **All Other Spanish/Hispanic/Latino** | 3037243 | 190321 (6.27%) | 2587336 (85.19%) | 68300 (2.25%) | 191286 (6.30%) |

note:

|  |  |
| --- | --- |
| **% total Hispanic** | 18.84% |
| **% total non-Hispanic** | 81.16% |

<sup>a</sup>N/A refers to individuals who lived in group quarters or housing units that were vacant at the time of data collection
<sup>b</sup>service refers to paying a cell phone company or internet service provider
<sup>c</sup>No responses are regarding to access at home; may not be representative of access to public internet sources
<sup>d</sup>Total Spanish/Hispanic/Latino includes all related origins below (Mexican, Puerto Rican, Cuban, Dominican, Costa Rican, Guatemalan, Honduran, Nicaraguan, Panamanian, Salvadoran, Other Central American, Argentinean, Bolivian, Chilean, Colombian, Ecuadorian, Paraguayan, Peruvian, Uruguayan, Venezuelan, Other South American, Spaniard, All other Spanish/Hispanic/Latino)

**Figure S1.**
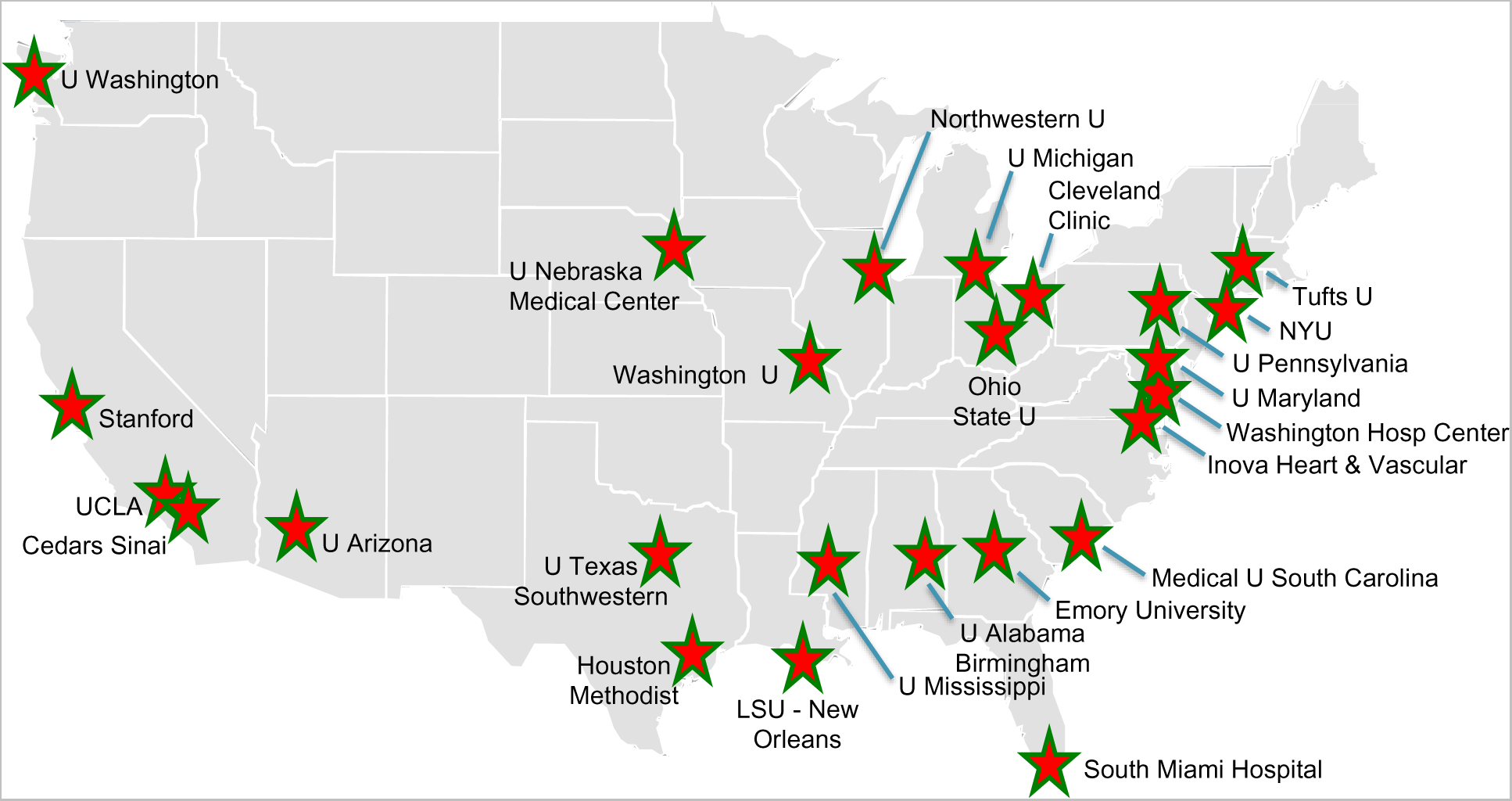
Clinical Sites of The DCM Consortium for the DCM Precision Medicine Study. Shown are the 25 clinical sites of the DCM Consortium who enrolled probands and family members for the DCM Precision Medicine Study. The Ohio State University site served as an enrolling site as well as the coordinating center.

**Figure S2.**
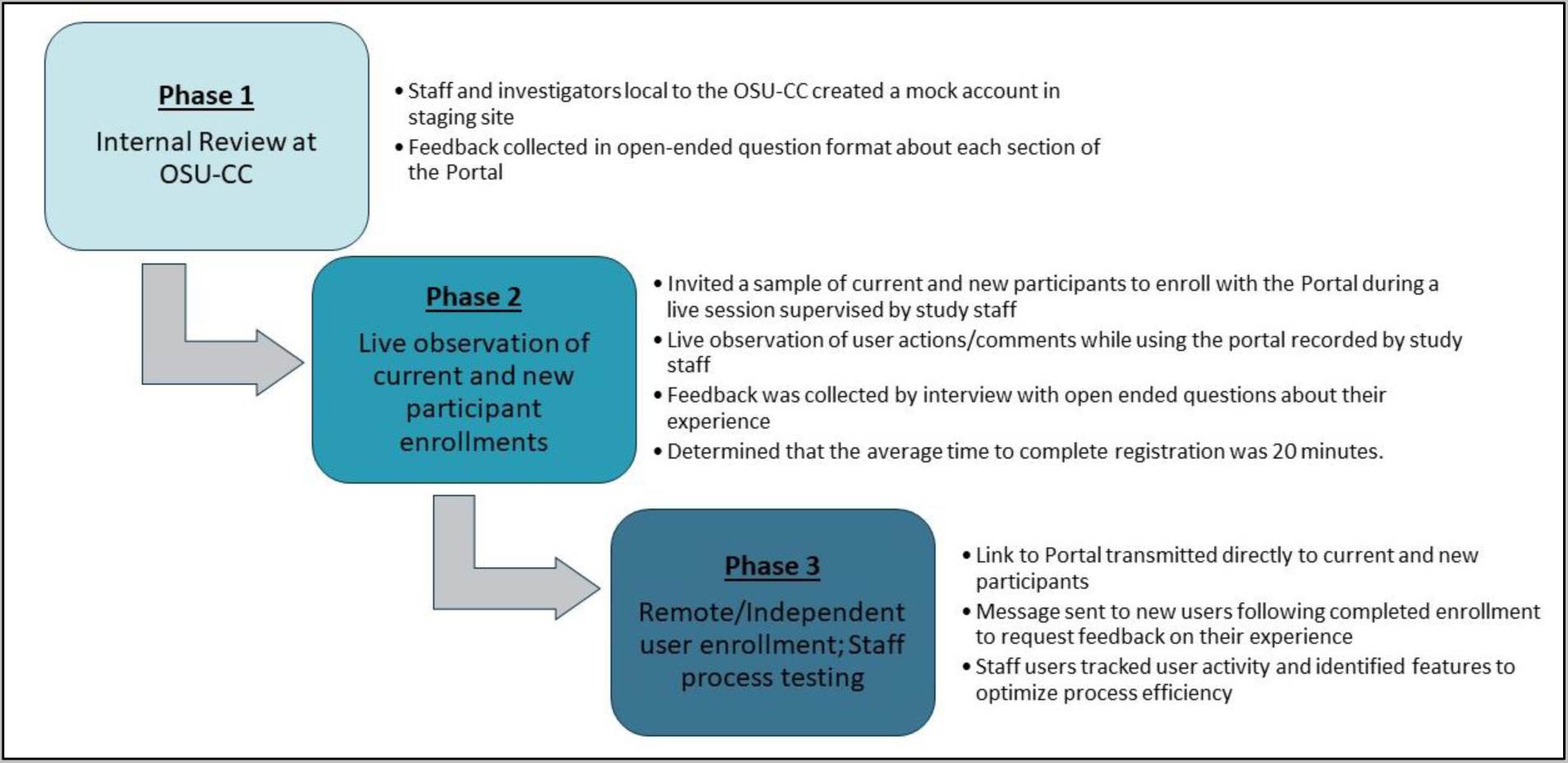
Beta testing phases of The DCM Project Portal. Figure S2. Three phases of beta testing were conducted before opening the DCM Project Portal for active service. Phase 1 included an internal review of staff and investigators familiar with the research project. Staff and investigators created mock accounts and offered feedback in an open-ended question form. After incorporating feedback, Phase 2 included a sample of participant users, including those previously consented to the study as well as newly invited. Individuals were observed going through the portal process live while a staff took notes of the actions and comments made during the experience. Feedback was collected directly once the process was completed by structured interview by the observing study staff member. Adjustments were made according to Phase 2 results before beginning Phase 3, where select participants (new and previously consented) were invited to create an account but this time independently. Progress was tracked by staff and feedback collected through the messaging system upon completion. Phase 3 findings included requests from both users and staff to maximize usability and efficiency.

